# SARS-CoV-2 vaccine breakthrough by Omicron and Delta variants: comparative assessments with New York State genomic surveillance data

**DOI:** 10.1101/2022.06.24.22276709

**Authors:** Alexander C. Keyel, Alexis Russell, Jonathan Plitnick, Jemma V. Rowlands, Eli Rosenberg, Daryl M. Lamson, Kathleen A. McDonough, Kirsten St. George

## Abstract

**Background:** Recently emerged variants of SARS-CoV-2 have shown greater potential to cause vaccine breakthrough infections.

**Methods:** A matched cohort analysis used a genomic sequence dataset linked with demographic and vaccination information from New York State (NYS). Two sets of conditional logistic regression analyses were performed, one during the emergence of Delta and another during the emergence of Omicron. For each set, cases were defined as individuals with the emerging lineage, and controls were individuals infected with any other lineage. The adjusted associations of vaccination status, vaccine type, time since vaccination, and age with lineage were assessed using odds ratios (OR) and 95% confidence intervals (CI).

**Results:** Fully vaccinated status (OR: 3, 95% CI: 2.0 - 4.9) and Boosted status (OR 6.7, 95% CI: 3.4 – 13.0) were significantly associated with having the Omicron lineage during the Omicron emergence period. Risk of Omicron infection relative to Delta generally decreased with increasing age (OR: 0.964, 95% CI 0.950 – 0.978). The Delta emergence analysis had low statistical power for the observed effect size.

**Conclusions:** Vaccines offered less protection against Omicron, thereby increasing the number of potential hosts for the emerging variant.

**Lay Summary:** There are different variants, or types, of the virus that causes COVID-19. These variants may differ in their ability to infect a person, cause severe disease, or evade vaccine protection. From previous studies, we know that vaccines provide substantial protection against the original COVID-19 virus. In this study, we wanted to know how some of the new variants compare to one another in this regard. We found that the Omicron variant could break through vaccine protection more effectively than the Delta variant. The data suggested that Delta may be better able to break through vaccines compared to previous variants, including Alpha, but our sample sizes were low, so this pattern was not statistically significant.

Individuals with a booster shot had much stronger protection against Delta compared to their protection against Omicron. We also found that younger people were more likely to be infected with Omicron than Delta.

## Introduction

As of June 1, 2022, the SARS-CoV-2 pandemic claimed >6.2 million human lives globally, with >1 million deaths in the United States (US) and >68,000 in New York State (NYS) [1]. SARS-CoV-2 has led to substantial agricultural losses [2], poses a risk to wildlife [3,4], and has spilled over into animal populations such as deer [5]. Viral evolution and adaptation has been observed in persistently-infected immunocompromised individuals [6] and animal reservoirs [7].

Novel variants of SARS-CoV-2 have shown increased rates of transmission and immune evasion [8,9]. In particular, Omicron has evolved a suite of unique mutations which have greatly increased its infectiousness [10], increased its ability to evade current vaccines [8,9], and decreased the effectiveness of convalescent plasma transfusions and monoclonal antibody treatments [11,12]. To a lesser degree, the Delta variant showed some of these same patterns of increased infectiousness [13] and potential for immune evasion compared to earlier strains [14].

Prior literature has also shown differences in vaccine effectiveness for SARS-CoV-2 lineages associated with variation in vaccine type, time post-vaccination, and patient age. Prior to Delta and Omicron, data have shown reduced neutralizing antibody protection for the Janssen vaccine compared to the Pfizer and Moderna vaccines [15] and slightly stronger protection for Moderna compared to Pfizer [15]. An effect of time post-vaccination has been demonstrated for the Delta variant [14]. Younger individuals were found to be more likely to be infected with Omicron [16,17].

A matched cohort study was used to test the associations of vaccination status, vaccine type, and time since vaccination with lineage identity during the emergence of new variants of SARS-CoV-2. Analyses were performed for the emergence of the Omicron and Delta variants in NYS.

## Methods

### Data Analysis

#### **Omicron Emergence Analysis (**28 November 2021 and 24 January 2022, Fig. 1)

Case-control matching (general approach): In this study design, individuals with Omicron (cases) were matched to individuals with any other lineage (controls). Cases (n = 1439) included B.1.1.529 and all BA sublineages (note: none were classified as BA.2 and BA.3 at the time of the analysis). Controls (n = 728) were all other lineages circulating during the period of Omicron emergence (all sequenced control samples in the matched data set were Delta variant: B.1.617.2 or AY sublineages). The start of the Omicron emergence period was defined by the first detection in the genomic surveillance dataset (even though Omicron was present in the state prior to this date). The emergence period ended when the last non-Omicron case was detected in the surveillance data set. Note that one additional Delta case was identified > 14 days after the last date in the surveillance data set but was excluded on the basis that the sensitivity analysis indicated it would not substantively change the analysis results. Cases were matched to controls on the basis of specimen collection date (±6 days), economic region (see Fig. 1), age, and sex (Male, Female). Age was matched according to bins (0 – 4, 5 – 11, 12 – 17, 18 – 29, 30 – 49, 50 – 69, 70 – 89, >90 years). If an exact match could not be found, mismatches were allowed for sex. One-to-one matching was used, without replacement (i.e., each case was matched to a unique control). The matching was performed in two stages. In the first stage, all possible matches were considered for each case. Cases were then sorted such that the cases with the fewest possible matches would be matched to controls first, in an effort to maximize the sample size.

**Fig. 1.**
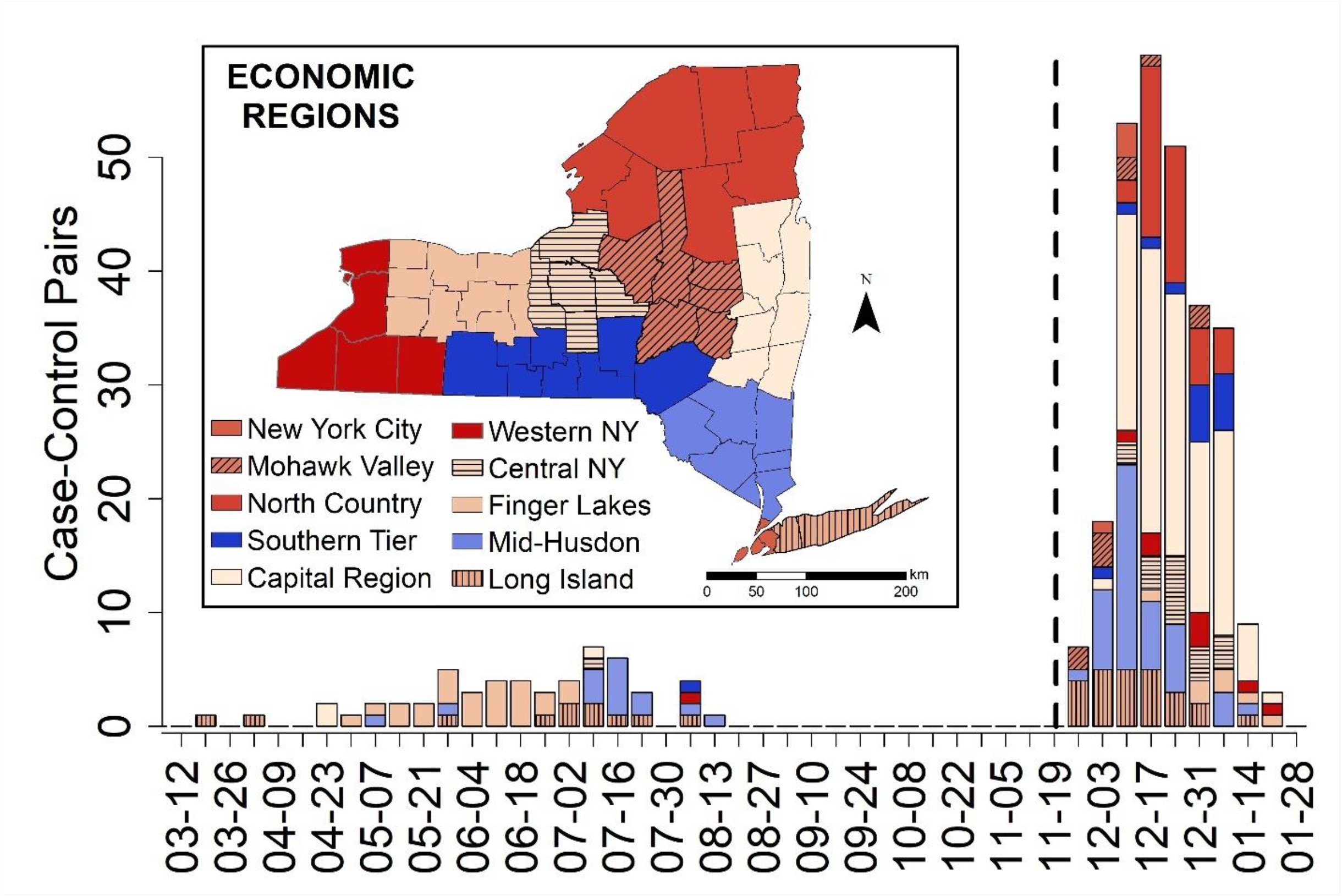
The number of matched case-control pairs used in the Conditional Logistic Regression by analysis for Delta (19 March 2021 – 15 August 2021) and Omicron (28 November 2021 −24 January 2022) emergence periods by New York State Economic Region (see inset). The bars correspond to the order given in the legend, with New York City on top when present, and Long Island on bottom when present. The dashed line was placed to separate the two data sets used in the analyses, with the Delta emergence period on the left and the Omicron emergence period on the right.

Three sets of conditional logistic regressions were performed to estimate Odds Ratios (OR) and 95% confidence intervals (CI). In the first analysis, both vaccinated and unvaccinated individuals were included. Key variables tested were vaccination status (binary: yes/no), booster status (yes/no), vaccine type (none, Pfizer, Moderna, Janssen), time since last vaccination or booster (three factor levels: unvaccinated, vaccinated <90 days, vaccinated >= 90 days). Time since completion of initial vaccination and time since booster were explored but were less predictive and overlapped strongly with the combined time since last vaccination or booster variable and therefore were excluded.

For the second analysis, the association between age and lineage was examined, and therefore age was removed as a matching criterion. A conditional logistic regression was performed using age, and interactions of age with the other main variables. No sorting prior to matching was performed for this analysis.

In the third analysis, cases were again matched to controls based on age, but unvaccinated individuals were excluded in order to allow time since last dose (vaccination series or booster) to be treated as continuous variables. Unvaccinated individuals could not be included in this analysis, as assigning them “NA” would cause these values to be excluded, and 0 would cause these cases to have an unrealistic value.

Finally, the leverage for each individual data point was tested by removing each case-control pair sequentially, refitting the model and noting the change in the odds ratio (OR). Models were selected using Akaike Information Criterion (AIC) scores [18,19]. Models with lower AIC scores have more model support, and models with ΔAIC >2 are generally considered less likely models. As a more complicated nested model can be within ΔAIC of 2, nested models were required to be within 2*K to be considered tied [20]. Note that AIC provides a relative ranking of models but provides no information on the absolute fit of the model. The fit of each model was examined by considering its statistical significance and the OR estimates. Where test results were not significant, the magnitude of the OR was examined. More research was deemed necessary if the estimated OR was large enough to be a public health concern, but 95% confidence intervals included 1.

All analyses were performed in R 4.1.2 [21] using the package survival for conditional logistic regressions [22,23]. R code is available from www.github.com/akeyel/CLR. The NYS map was created in ArcGIS 10.6 (ESRI, Redlands, CA) using a 2017 Tiger Shapefile from the US Census Bureau [24] and Admin 1 States, provinces 50-m-cultural vector shapefile from Natural Earth Data (accessed 18 March 2022) (https://www.naturalearthdata.com/downloads/50m-cultural-vectors/)

#### **Delta Emergence Analysis (**19 March 2021 and 15 August 2021, Fig. 1)

The Delta analyses followed the same methods described above for the Omicron analyses. For the Delta analyses, the focal lineages (cases, n = 603) included B.1.617.2 and all AY sublineages. Non-focal lineages (controls, n = 1816) were all other lineages circulating during the period of Delta emergence (62% B.1.1.7 and Q.4 Alpha, 20% B.1.526 Iota, 3.5% P.1.X Gamma, 1% B.1.351.X Beta, none of the other non-Variant of Concern strains (13.7% combined) exceeded 5%. Booster-related variables were excluded from the analysis as booster shots were not widely available during this period. No analysis was performed on vaccinated only individuals due to low statistical power resulting from a sample size of 12 matched pairs.

### Power Analysis

Statistical power for conditional logistic regression is non-linear, and depends on estimated probabilities. While the analyses above used multiple conditional logistic regression, statistical power was calculated for univariate logistic regression using the WebPower package [25,26] as a simplifying assumption, in order to make the power analyses easier to set up and interpret. Statistical power to detect an OR of 2 with a sample size of 110 was examined for a range of probability values (0.1 – 0.9 for the upper probability, lower probability was adjusted to give an OR of 2). The upper probability value with the highest power (0.7) was then used to assess statistical power for ORs of 2, 3, and 4 for sample sizes between 50 and 350 by increments of 50.

### Data Sources

Respiratory swab specimens, positive for SARS-CoV-2 by real-time RT-PCR, were sent from clinical laboratories across NYS for whole genome sequencing at the NYSDOH Wadsworth Center as part of an enhanced genomic surveillance program. Samples were matched to demographics in the Communicable Disease Electronic Surveillance System (CDESS) and vaccination records in the New York State Immunization System (NYSIIS). For persons with multiple collections, only the earliest collected sample with genome available was included.

Vaccination status was determined for each individual based on dates of sample collection and administration of vaccines. A person was considered “unvaccinated” if the sample was collected prior to any vaccination, “vaccinated” if the sample was collected >14 days after completion of vaccination (first shot of Janssen, second shot of Pfizer or Moderna), and “boosted” if the sample was collected any time after receiving a booster dose, of any vaccine type. Individuals with partial vaccination (sample collected between initial dose and 14 days post completion of vaccination) and those who received a greater number of vaccinations than normal were removed from the study. This study does not apply to individuals who received a 3^rd^ dose as part of their vaccination series (e.g., potentially immunocompromised individuals), as these individuals were removed from the data set (due to different vaccination history and low sample sizes, 58 individuals removed with a 3^rd^ shot less than 135 days from their second shot).

### Sequencing Methods

Whole genome amplicon sequencing of SARS-CoV-2 was performed using a modified version of the Illumina ARTIC protocol (https://artic.network/ncov-2019) using ARTIC V3 primers in the Applied Genomics Technology Core at the Wadsworth Center, as previously described [27], with later samples amplified with ARTIC V4 primers.

For samples of particularly low viral titer, sequencing was performed using the Ion Torrent S5XL sequencer, as previously described **[28]**. GISAID Accession Sequences are available from www.github.com/akeyel/CLR/GISAID_accession_IDs.csv The first column includes the GISAID accession ID. The subsequent columns indicate whether the ID was used in the respective analyses. Data are coded such that -1 indicates records that were removed prior to analysis, 0 indicates records that met the basic overall study criteria, but that were not matched for a particular analysis, and 1 indicates that the record was included in the analysis.

## Results

### Omicron Emergence

In Analysis 1, >80% of case/control pairs were aged 18 – 69, with the majority from the Capital Region and Mid-Hudson (Table 1, Fig. 1). 8% of control individuals had received a booster while 22% of cases had been boosted. 56.6% of controls and 30% of cases were unvaccinated (Table 1). Sample sizes were 177 for Pfizer, 109 for Moderna, and 22 for Janssen. Vaccination (OR 3.1, CI 2.0 – 4.9, p<0.001) and booster status (OR 6.7, CI 3.4 – 13.0, p <0.001) were the variables most associated with an Omicron lineage identity (Table 2, Fig. 2).

**Table 1.** Descriptive statistics for matched cases and controls for the Conditional Logistic Regression for the Omicron emergence period. Presence (case) or absence (control) of Omicron was used as the basis for matching.

|  | Analysis 1 (main) |  |  |  | Analysis 2 (age) |  |  |  | Analysis 3 (vax only) |  |  |  |
| --- | --- | --- | --- | --- | --- | --- | --- | --- | --- | --- | --- | --- |
| Demographic group | Control | % | Case | % | Control | % | Case | % | Control | % | Case | % |
|  | Count |  | Count |  | Count |  | Count |  | Count |  | Count |  |
| **Age** |  |  |  |  |  |  |  |  |  |  |  |  |
| 00-04 | 4 | 1.5 | 4 | 1.5 | 9 | 2.9 | 4 | 1.3 | 0 | 0 | 0 | 0 |
| 05-11 | 4 | 1.5 | 4 | 1.5 | 7 | 2.3 | 9 | 2.9 | 0 | 0 | 0 | 0 |
| 12-17 | 11 | 4 | 11 | 4 | 15 | 4.9 | 16 | 5.2 | 3 | 2.3 | 3 | 2.3 |
| 18-29 | 55 | 20.2 | 55 | 20.2 | 39 | 12.6 | 85 | 27.5 | 23 | 17.8 | 23 | 17.8 |
| 30-49 | 95 | 34.9 | 95 | 34.9 | 85 | 27.5 | 95 | 30.7 | 49 | 38 | 49 | 38 |
| 50-69 | 71 | 26.1 | 71 | 26.1 | 96 | 31.1 | 69 | 22.3 | 40 | 31 | 40 | 31 |
| 70-89 | 31 | 11.4 | 31 | 11.4 | 52 | 16.8 | 30 | 9.7 | 14 | 10.9 | 14 | 10.9 |
| ≥90 | 1 | 0.4 | 1 | 0.4 | 6 | 1.9 | 1 | 0.3 | 0 | 0 | 0 | 0 |
| **Sex** |  |  |  |  |  |  |  |  |  |  |  |  |
| Male | 141 | 51.8 | 147 | 54 | 155 | 50.2 | 153 | 49.5 | 65 | 50.4 | 77 | 59.7 |
| Female | 129 | 47.4 | 123 | 45.2 | 152 | 49.2 | 155 | 50.2 | 63 | 48.8 | 52 | 40.3 |
| Unknown | 2 | 0.7 | 2 | 0.7 | 2 | 0.6 | 1 | 0.3 | 1 | 0.8 | 0 | 0 |

| **Region** |  |  |  |  |  |  |  |  |  |  |  |  |
| --- | --- | --- | --- | --- | --- | --- | --- | --- | --- | --- | --- | --- |
| Capital Region | 107 | 39.3 | 107 | 39.3 | 121 | 39.2 | 121 | 39.2 | 47 | 36.4 | 47 | 36.4 |
| Central New York | 17 | 6.2 | 17 | 6.2 | 18 | 5.8 | 18 | 5.8 | 10 | 7.8 | 10 | 7.8 |
| Finger Lakes | 7 | 2.6 | 7 | 2.6 | 9 | 2.9 | 9 | 2.9 | 3 | 2.3 | 3 | 2.3 |
| Long Island | 25 | 9.2 | 25 | 9.2 | 27 | 8.7 | 27 | 8.7 | 12 | 9.3 | 12 | 9.3 |
| Mid-Hudson | 42 | 15.4 | 42 | 15.4 | 47 | 15.2 | 47 | 15.2 | 26 | 20.2 | 26 | 20.2 |
| Mohawk Valley | 10 | 3.7 | 10 | 3.7 | 18 | 5.8 | 18 | 5.8 | 4 | 3.1 | 4 | 3.1 |
| New York City | 4 | 1.5 | 4 | 1.5 | 6 | 1.9 | 6 | 1.9 | 1 | 0.8 | 1 | 0.8 |
| North Country | 38 | 14 | 38 | 14 | 39 | 12.6 | 39 | 12.6 | 20 | 15.5 | 20 | 15.5 |
| Southern Tier | 14 | 5.1 | 14 | 5.1 | 16 | 5.2 | 16 | 5.2 | 2 | 1.6 | 2 | 1.6 |
| Western New York | 8 | 2.9 | 8 | 2.9 | 8 | 2.6 | 8 | 2.6 | 4 | 3.1 | 4 | 3.1 |
| **Vaccination Status** |  |  |  |  |  |  |  |  |  |  |  |  |
| Unvaccinated | 154 | 56.6 | 82 | 30.1 | 175 | 56.6 | 78 | 25.2 | 0 | 0 | 0 | 0 |
| Vaccinated <90 days | 3 | 1.1 | 4 | 1.5 | 3 | 1 | 5 | 1.6 | 2 | 1.6 | 2 | 1.6 |
| Vaccinated >90 days | 115 | 42.3 | 186 | 68.4 | 131 | 42.4 | 226 | 73.1 | 127 | 98.4 | 127 | 98.4 |
| Pfizer Vaccine | 64 | 23.5 | 113 | 41.5 | 69 | 22.3 | 135 | 43.7 | 64 | 49.6 | 82 | 63.6 |
| Moderna Vaccine | 43 | 15.8 | 66 | 24.3 | 49 | 15.9 | 82 | 26.5 | 48 | 37.2 | 41 | 31.8 |
| Janssen Vaccine | 11 | 4 | 11 | 4 | 16 | 5.2 | 14 | 4.5 | 17 | 13.2 | 6 | 4.7 |
| Unboosted | 250 | 91.9 | 211 | 77.6 | 281 | 90.9 | 210 | 68 | 108 | 83.7 | 88 | 68.2 |
| Boosted <90 days | 18 | 6.6 | 49 | 18 | 25 | 8.1 | 76 | 24.6 | 18 | 14 | 37 | 28.7 |
| Boosted >90 days | 2 | 0.7 | 9 | 3.3 | 2 | 0.6 | 20 | 6.5 | 1 | 0.8 | 3 | 2.3 |
| Pfizer Booster | 11 | 4.8 | 41 | 15.4 | 13 | 4.5 | 68 | 22.7 | 10 | 7.8 | 33 | 25.6 |
| Moderna Booster | 9 | 3.3 | 17 | 7 | 14 | 4.5 | 28 | 9.4 | 9 | 7 | 7 | 5.4 |

**Table 2.** Odds ratios (95% confidence intervals) and parameter significance for models of particular interest for the three Omicron analyses. ^1^ There was only one boosted individual with an initial Janssen shot, so for statistical reasons this individual was pooled with the unboosted Janssen individuals. When fit separately, the odds ratio for unboosted Janssen individuals was 1.9 (0.8 – 4.9) with a parameter p value of 0.20, and the parameter estimate for the single boosted Janssen individual was unreliable.

| Model | ΔAI | Parameter 1 | Parameter 2 | Parameter 3 | | |
| --- | --- | --- | --- | --- | --- | --- |
|  | C |  |  |  |  |  |
| **Main Analysis** |  |  |  |  |  |  |
| Vaccination Status | 0.00 | Vaccinated | Vaccinated +<br>Boosted |  |  |  |
|  |  | 3.1 (2.0 -4.9) *** | 6.7 (3.4 –<br>13.0)*** |  |  |  |
| Vaccine Type | 3.33 | Pfizer | Moderna | Janssen<sup>1</sup> |  |  |
| without booster: |  | 3.3 (1.9 - 5.6)*** | 3.6 (2.0 – 6.7)*** | 2.0 (0.8 – 5.1) |  |  |
| with booster |  | 10.4 (4.3 – 25.2)*** | 3.8 (1.5 – 9.3)** |  |  |  |
| **Age Analysis** |  |  |  |  |  |  |
| Vaccination Status +<br>Age | 0.00 | Vaccinated | Vaccinated +<br>Boosted | Age<br>(linear) | Age<br>(0 - 4) | Age<br>(18 - 29) |
| + Age bins |  |  |  |  |  |  |
|  |  | 4.8 (2.8 – 8.1)*** | 38.5 (15.9 – 93.2) | 0.964 | 0.250 | 2.0 |
|  |  |  | *** | (0.950- | (0.059- | (1.1- |
|  |  |  |  | 0.978) | 1.051) | 3.7) |
|  |  |  |  | *** |  | * |
| Vaccination Status +<br>Age | 7.41 | Vaccinated | Vaccinated +<br>Boosted | Age |  |  |
|  |  | 5.0 (3.0 – 8.3)*** | 34.1 (14.6 – 79.5) | 0.962 (0.950 - 0.974)*** |  |  |
|  |  |  | *** |  |  |  |
| **Vax Only Analysis** |  |  |  |  |  |  |
| Janssen + Days Post Dose | 0.00 | Janssen (relative to<br>mRNA vaccine) |  | Days post last dose (booster<br>or primary series) |  |  |
|  |  | 0.388 (0.149 –<br>1.009) |  | 0.996 (0.993 – 0.999)** |  |  |
| Vaccine Type +<br>Days Post Dose | 1.18 | Moderna (relative to<br>Pfizer) | Janssen (relative<br>to Pfizer) | Days Post last dose (booster<br>or primary series) |  |  |
|  |  | 0.776 (0.448 –<br>1.344) | 0.351 (0.132-0.935)* | 0.996 (0.993 – 0.999)** |  |  |
| Parameter p-values: * p < 0.05, ** p < 0.01, *** p < 0.001 |  |  |  |  |  |  |

**Fig. 2.**
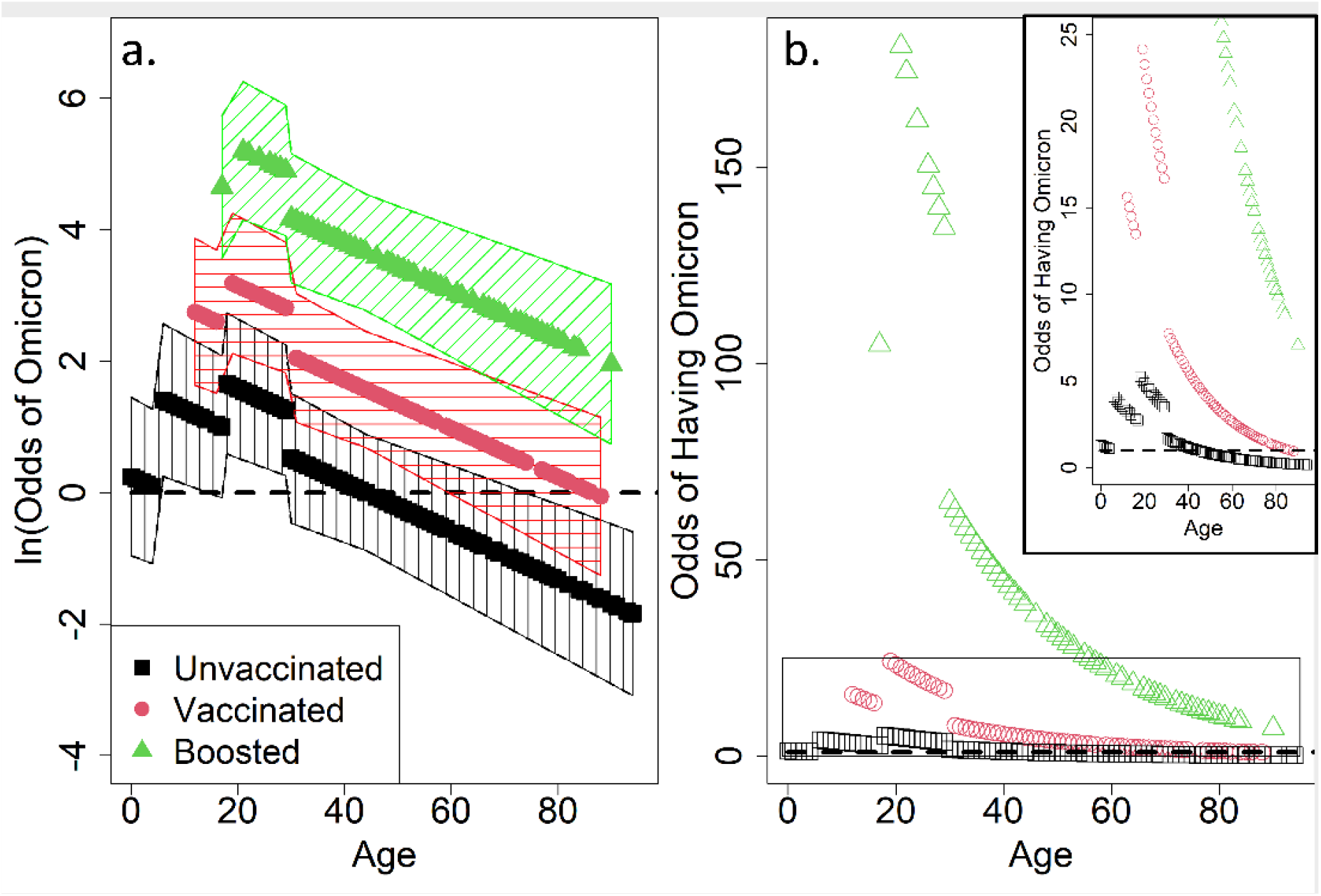
Visualization of the fixed effects (excluding stratum-specific effects, which were often strong) from the Omicron Emergence analysis without age matching for the data set used to generate the analysis a) on a log-odds scale and b) on an odds scale. The inset shows Odds from 0 – 25 with greater resolution. Odds of 1 or Log(Odds) of 0 indicate an equal probability of having Omicron or Delta, with increasing values indicative of an increasing probability of having Omicron instead of Delta. Hatched areas indicate +/- 1 SE, and are only shown in (a) for clarity.

In Analysis 2, when age was removed as a matching criterion, younger age was also predictive of an Omicron infection, with odds of having Omicron generally decreasing as age increased (OR 0.962, CI 0.950 – 0.974, Table 2). Individuals 0-4 years of age had a lower risk of Omicron than predicted by a log-linear age effect (but still much higher risk compared to the 90+ age group), while individuals 18 – 29 had a higher risk of Omicron than predicted by a log-linear age term, and the highest risk of any age group. Estimates for OR for vaccination status (OR 4.8, CI 2.8 – 8.1) and booster status (OR 38.5, CI 15.9 – 93.2) were higher than in in the analysis that matched on age (Table 2).

In Analysis 3, when only vaccinated individuals were considered, the probability of having Omicron decreased with an increase in the number of days following the last vaccine dose (OR 0.996, CI 0.993 - 0.999, Table 2). Vaccine type was also included in the top statistical models, with a borderline significant trend towards reduced odds of Omicron with the Janssen vaccine (OR 0.351, CI: 0.132 - 0.935, relative to Pfizer vaccine; OR 0.388, CI 0.149 - 1.009, relative to any mRNA vaccine, Table 2). AIC scores for alternative statistical models are presented in Table S1.

### Delta Emergence

In Analysis 1, 75% of case/control pairs were aged 18 – 69, with 89% of case/controls from Finger Lakes, Long Island, and the Mid-Hudson regions (Table 3). 74.5% of controls and 61.8% of cases were unvaccinated (Table 3). Neither vaccine type, time from last vaccination, nor an interaction of the two were significantly related to an increased likelihood of having Delta than any other lineage in the fully matched conditional logistic regression (Table 4). Vaccination status was the ‘best’ model (OR 2.4, CI 0.8 – 6.8, p = 0.08, Table 4). There was no significant effect of vaccine type (p = 0.12), but detected ORs were 2.9 (Pfizer, CI: 0.9 – 8.9), 0.38 (Moderna, CI: 0.04 – 4.2), and 2.0 (Janssen, CI: 0.17 – 23.6).

**Table 3.**
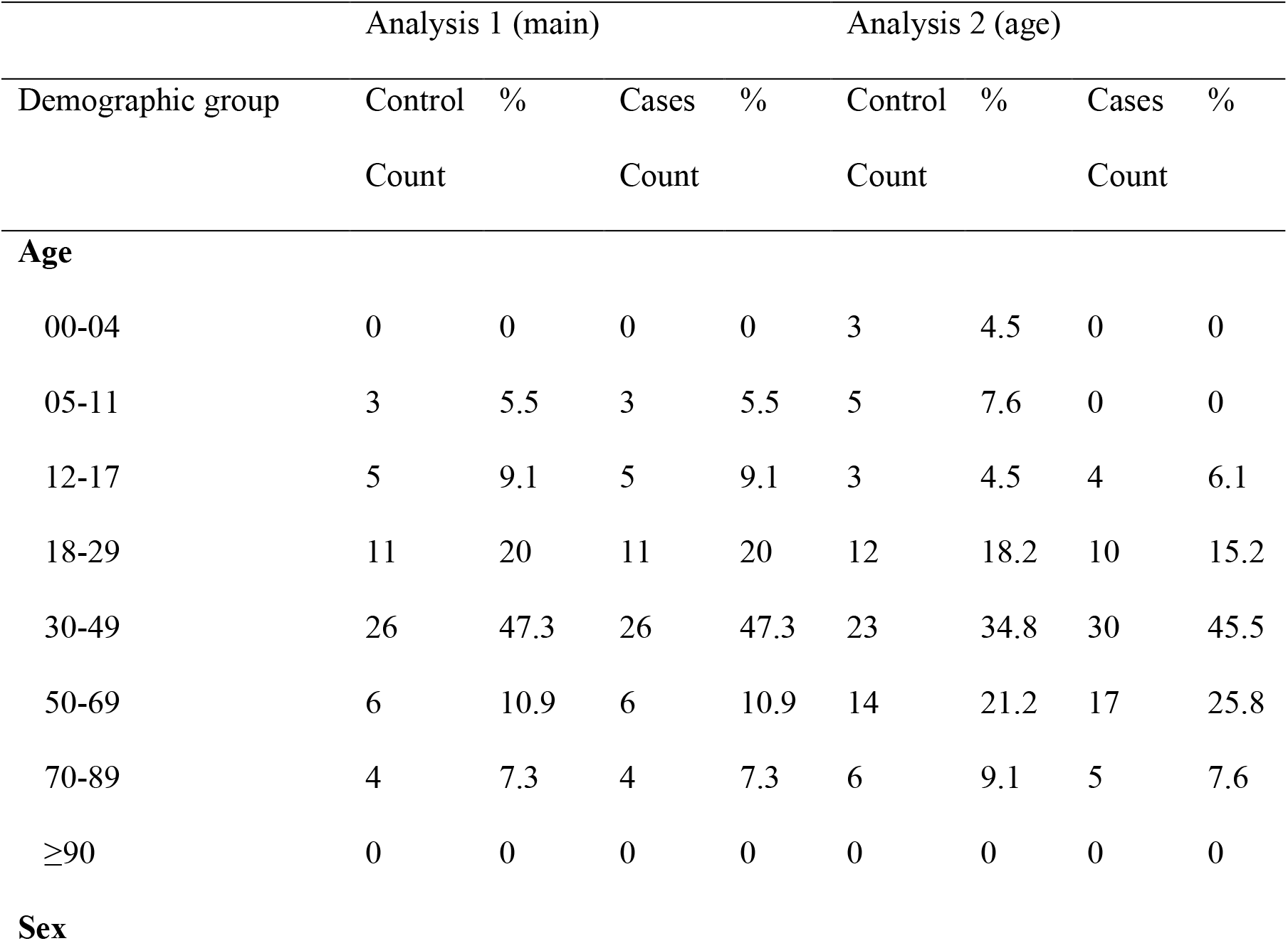
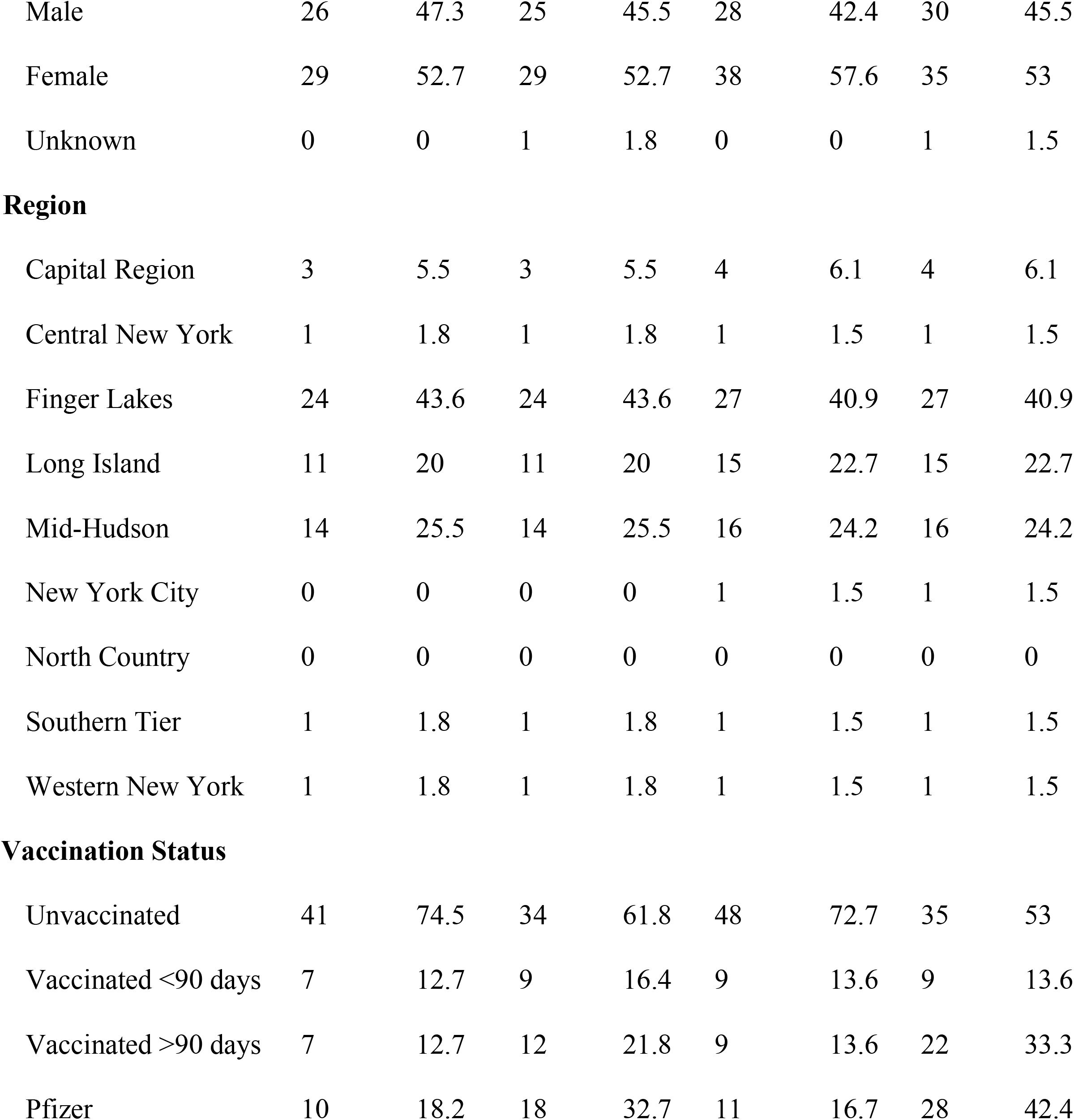
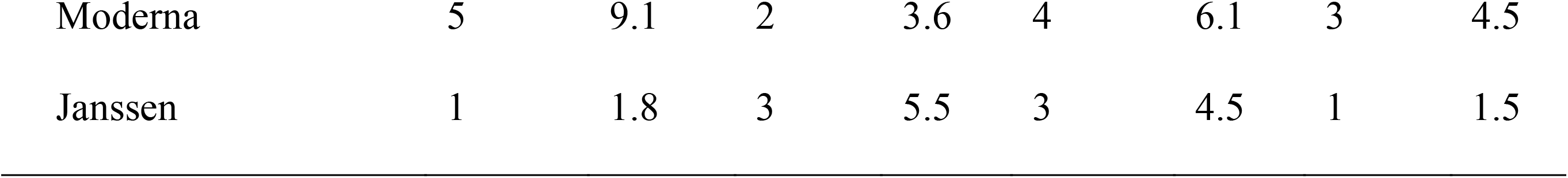
Descriptive statistics for matched cases and controls for the Conditional Logistic Regression for the Delta emergence period. Presence (case) or absence (control) of Delta was used as the basis for matching.

|  | Analysis 1 (main) |  |  |  | Analysis 2 (age) |  |  |  |
| --- | --- | --- | --- | --- | --- | --- | --- | --- |
| Demographic group | Control | % | Cases | % | Control | % | Cases | % |
|  | Count |  | Count |  | Count |  | Count |  |
| **Age** |  |  |  |  |  |  |  |  |
| 00-04 | 0 | 0 | 0 | 0 | 3 | 4.5 | 0 | 0 |
| 05-11 | 3 | 5.5 | 3 | 5.5 | 5 | 7.6 | 0 | 0 |
| 12-17 | 5 | 9.1 | 5 | 9.1 | 3 | 4.5 | 4 | 6.1 |
| 18-29 | 11 | 20 | 11 | 20 | 12 | 18.2 | 10 | 15.2 |
| 30-49 | 26 | 47.3 | 26 | 47.3 | 23 | 34.8 | 30 | 45.5 |
| 50-69 | 6 | 10.9 | 6 | 10.9 | 14 | 21.2 | 17 | 25.8 |
| 70-89 | 4 | 7.3 | 4 | 7.3 | 6 | 9.1 | 5 | 7.6 |
| ≥90 | 0 | 0 | 0 | 0 | 0 | 0 | 0 | 0 |
| **Sex** |  |  |  |  |  |  |  |  |
| Male | 26 | 47.3 | 25 | 45.5 | 28 | 42.4 | 30 | 45.5 |
| Female | 29 | 52.7 | 29 | 52.7 | 38 | 57.6 | 35 | 53 |
| Unknown | 0 | 0 | 1 | 1.8 | 0 | 0 | 1 | 1.5 |
| **Region** |  |  |  |  |  |  |  |  |
| Capital Region | 3 | 5.5 | 3 | 5.5 | 4 | 6.1 | 4 | 6.1 |
| Central New York | 1 | 1.8 | 1 | 1.8 | 1 | 1.5 | 1 | 1.5 |
| Finger Lakes | 24 | 43.6 | 24 | 43.6 | 27 | 40.9 | 27 | 40.9 |
| Long Island | 11 | 20 | 11 | 20 | 15 | 22.7 | 15 | 22.7 |
| Mid-Hudson | 14 | 25.5 | 14 | 25.5 | 16 | 24.2 | 16 | 24.2 |
| New York City | 0 | 0 | 0 | 0 | 1 | 1.5 | 1 | 1.5 |
| North Country | 0 | 0 | 0 | 0 | 0 | 0 | 0 | 0 |
| Southern Tier | 1 | 1.8 | 1 | 1.8 | 1 | 1.5 | 1 | 1.5 |
| Western New York | 1 | 1.8 | 1 | 1.8 | 1 | 1.5 | 1 | 1.5 |
| **Vaccination Status** |  |  |  |  |  |  |  |  |
| Unvaccinated | 41 | 74.5 | 34 | 61.8 | 48 | 72.7 | 35 | 53 |
| Vaccinated <90 days | 7 | 12.7 | 9 | 16.4 | 9 | 13.6 | 9 | 13.6 |
| Vaccinated >90 days | 7 | 12.7 | 12 | 21.8 | 9 | 13.6 | 22 | 33.3 |
| Pfizer | 10 | 18.2 | 18 | 32.7 | 11 | 16.7 | 28 | 42.4 |
| Moderna | 5 | 9.1 | 2 | 3.6 | 4 | 6.1 | 3 | 4.5 |
| Janssen | 1 | 1.8 | 3 | 5.5 | 3 | 4.5 | 1 | 1.5 |

**Table 4.** Odds Ratios (95% confidence intervals) and parameter significance for selected models from the two Delta analyses. Note that none of the main analysis models were statistically significant, as all 95% confidence intervals for odds ratio estimates overlapped 1.

| Model | ΔAI | Parameter 1 | Parameter 2 | Parameter 3 |
| --- | --- | --- | --- | --- |
| C |  |  |  |  |
| **Main Analysis** |  |  |  |  |
| Vaccination Status | 0.00 | Vaccinated |  |  |
|  |  | 2.4 (0.8 -6.8) |  |  |
| Vaccine Type | 1.05 | Pfizer | Moderna | Janssen |
|  |  | 2.86 (0.92 – 8.94) | 0.38 (0.04 – 4.20) | 1.97 (0.17 – 23.57) |
| **Age Analysis** |  |  |  |  |
| Vaccine Type | 0.02 | Pfizer | Moderna | Janssen |
|  |  | 7.3 (2.0 – 26.7)** | 2.0 (0.25 – 17.1) | 0.46 (0.04 – 4.76) |
| Parameter p-values: * $p < 0.05$, ** $p < 0.01$, *** $p < 0.001$ | | | | |

The power analysis showed that a sample size of 110 (55 pairs) would have a 15 - 45% chance of obtaining a significant result for an OR of 2 under the simulated probability distributions. A sample size of at least 255 would be needed to have ≥80% power for an OR of 2. A sample size of 110 could have up to 78% power to detect an OR of 3 and 93% power for an OR of 4. A sample size of 24 could detect an OR of 22 with 80% power, but would only have 36% power to detect an OR of 4.

When cases and controls were no longer matched on the basis of age, vaccine type was the ‘best’ model, suggesting that odds of having Delta rather than any other lineage increased by a factor of 7.3 (2.0 - 26.7) for those receiving the Pfizer vaccine relative to unvaccinated individuals. Effects for Moderna (2.0, 95% CI: 0.25 - 17.1) and Janssen (0.46, 95% CI: 0.04 - 4.76) were substantial but not individually significant. When the analysis was restricted to only vaccinated individuals, the sample size was too small for meaningful analysis (12 matched pairs). AIC scores for alternative statistical models are presented in Table S2.

## Discussion

This study adds to the body of evidence supporting immune escape of SARS-CoV-2 by exploring vaccine breakthrough, vaccination status, and time since vaccination in a matched case-control cohort study. As a consequence of the study design, some of the results may seem counter intuitive. For example, while a booster increases protection against Omicron compared to an absence of a booster [16,29], history of a booster was associated with Omicron and not Delta. This is consistent with evidence that suggests that having a booster is less effective for preventing infection with Omicron than with Delta [9,16]. Similarly, vaccine effectiveness has been shown to wane with time [14], therefore it was hypothesized that increased time following vaccination would decrease the odds of being infected with the emergent strain.

Our analysis of NYS genomic surveillance data yielded results that are consistent with previous research showing an increased probability of breakthrough for Omicron compared to other variants for both vaccinated and boosted individuals [9,11]. In a similar study in Connecticut comparing odds of Omicron vs. Delta infection [9], an OR of ∼2 (CI 1.5 - 3.7 or 1.5 - 2.2 depending on time post vaccination) was found for vaccinated individuals and ∼3 (1.8 - 4.9) for boosted individuals. These estimates are lower than the estimates from this study of 3.1 (2.0 - 4.9) for vaccinated individuals and 6.7 (3.4 - 13.0) for unvaccinated, but the 95% CIs overlap between the two studies. There is a strong pattern of the emergent strain showing increased ability to breakthrough vaccines compared to other strains circulating at the time. Studies on prior variants of concern have found significant vaccine breakthrough in emergent variants. For example, Kustin et al. 2021 found that Alpha (B.1.1.7) was more likely to show vaccine breakthrough compared to prior strains [30]. Similarly, Tartof et al. 2021 found evidence for increased rates of vaccine breakthrough in Delta (B.1.617.2), although waning vaccine immunity was also a factor in that study [14]. Similarly, Rosenberg et al. 2022 showed an increase in breakthroughs during the Delta emergence period, and suggested this effect existed independently of waning immunity [31].

Time post vaccination was a statistically significant factor in this study when the analysis was restricted to vaccinated individuals only, with probability of Omicron decreasing with increased time post vaccination. The time post vaccination variable combined individuals who were recently boosted with those who had recently completed their primary series. Adding a variable to indicate booster status did not improve the model fit (Table S1). It is notable that most individuals in this study were more than 3 months post completion of their initial vaccination series. Booster shots were generally more recent, and therefore vaccination status and booster status likely encoded much of the information that would have been conveyed by a time post last dose variable. No time post vaccination effect was detected if the data were coarsely divided into individuals with boosters and individuals without boosters, suggesting more examination of this variable may be necessary. This variable was not found among the top models in the Delta Emergence analysis.

Younger individuals were found to be more likely to be infected with Omicron than with Delta during the Omicron emergence period, although the data in this study cannot be used to distinguish a physiological basis from a behavioral basis for these age effects. Kahn et al. (2022) found equal age distributions for Delta and Omicron among unvaccinated individuals, but a strong shift towards younger individuals among those vaccinated (however see data table in Accorsi et al. 2022, in the US, where Omicron rates are elevated among both vaccinated and unvaccinated). It is possible that the age bin effects are due to a greater degree of socialization and other behavioral risk factors among 18 – 29 year-olds. For example, in 2020, college campus re-openings were associated with increased COVID transmission [32]. With Omicron’s ability to break through vaccinations, college campuses may have increased this age-group’s likelihood of being infected with SARS-CoV-2 (e.g., Wan et al. 2022). The bin effect for pre-school age children (0 – 4 years of age) may represent a reduced level of socialization for this group. This effect, while included in the ‘best’ model identified by the information theoretic approach here, was not statistically significant, so it also may be an artifact of low sample sizes in this age group. Other research has found that vaccines were not equally effective among age groups [33]. Vaccine effectiveness in NYS was very low for 5 – 11 year-olds who received a lower dose (10 μg) of the Pfizer vaccine than for vaccinated individuals aged 12 and up (30 μg dose) [33]. However, the log-linear age effect detected here was not driven by children under 12. When children under 12 were removed from the analysis, the estimated OR changed from 0.962 to 0.957 (CI 0.944 – 0.971), suggesting the magnitude of the effect is greater when young children were removed from the analysis. Larger estimates for vaccination status and booster status were also greater when children under 12 were removed from the analysis (vaccination status OR: 5.4, [CI: 3.1 – 9.7], vaccination plus booster status OR: 43.0 [CI: 17.1 – 108.5]).

Sample sizes were generally too small to detect robust vaccine type effects. The Janssen vaccine showed borderline significant reduced OR of Omicron relative to the Pfizer vaccine in one statistical model (Table 2, Table S1). This result would be consistent with improved performance against Omicron, or with worse performance of this vaccine against Delta, as has been observed [e.g., 31]. Otherwise, OR estimates showed the potential for substantial differences, but overlapping confidence intervals prevent drawing robust conclusions.

Low statistical power in this study was due to limited sample sizes, which were constrained by the limited emergence periods and the relatively small percentage of COVID-19 cases that are sequenced. For Delta, the emergence period occurred during a time of reduced sequencing, due to low overall incidence during the summer of 2021 when Delta displaced prior strains (Fig. 1). For Omicron, a larger sequencing effort was made, but the emergence period was considerably shorter due to the rapid dominance of the Omicron variant (Fig. 1). Sample sizes could potentially be increased by expanding the regional scope of the study or incorporating sequencing results from other research labs.

Only a single matched set was used for each analysis. However, cases were randomly matched to controls, and therefore other matches were possible. This limitation could be overcome by assessing significance with Monte Carlo simulation over the range of possible matches. That said, visual examination of leverage plots based on removing a single pair suggested that the results were generally unlikely to change with the removal of any single data point. The exception is the Delta analysis, where a change in 1-2 data points would change the overall statistical significance of the results (Fig. S1), without much change in the estimated OR.

## Data Availability

All de-identified data produced in the present study are available upon reasonable request to the authors, subject to NYSDOH approval.

https://www.github.com/akeyel/CLR

## Acknowledgements

The authors thank the Wadsworth Center’s Virology Laboratory for processing specimens, the Advanced Genomic Technologies Core for performing next-generation sequencing, and the Bioinformatics Core for sequence analysis. We also thank the New York State clinical laboratories that submitted SARS-CoV-2– positive specimens to Wadsworth for sequencing, as well as all originating and submitting laboratories for their SARS-CoV-2 sequence contributions to the GISAID database.

## Data Availability

The data set analyzed in this study is not publicly available.

---

**Table S1.** AIC Results for the Omicron Analyses. Results are for 3 separate conditional logistic regression analyses (see methods). For all analyses, each case of Omicron was matched to a non-Omicron control. Minimum AIC scores for each analysis were 330.38, 285.21, and 168.43, with corresponding log-likelihoods of -163.19, -137.61, and -82.21. Sample sizes were 272, 309, and 129 pairs. AIC scores SHOULD NOT be compared across different analyses. × indicates an interaction, while + indicates variables in a model without an interaction term. Full models did not include interaction effects. Best models are indicated in italics.

| Model | Delta<br>AIC | K | p-value |
| --- | --- | --- | --- |
| **1) Conditional Logistic Regression** |  |  |  |
| **(CLR)** |  |  |  |
| *Booster Status + Vaccination Status* | *0.00* | *2* | *<0.001\*\*\** |
| Time Post Dose | 2.37 | 2 | <0.001*** |
| Vaccination Status | 4.15 | 1 | <0.001*** |
| Vaccine Type | 5.46 | 3 | <0.001*** |
| Full Model | 6.19 | 6 | <0.001*** |
| Booster Status | 25.41 | 1 | <0.001*** |
| **2) No-age matching CLR** |  |  |  |
| *Age + Age Bin (18-29) + Age Bin (0 - 4) +* |  |  |  |
| *Vaccination Status + Booster Status* | *0.00* | *7* | *<0.001\*\*\** |
| Age + Vaccination Status + Booster Status | 7.41 | 3 | <0.001*** |
| Full Model | 11.73 | 7 | <0.001*** |
| Time Post Dose × Age | 29.29 | 5 | <0.001*** |
| Vaccine Type × Age | 43.35 | 7 | <0.001*** |
| Vaccination Status + Booster Status | 57.67 | 2 | <0.001*** |
| Age | 128.36 | 1 | **<0.001\*\*\*** |
| **3) Vaccinated-only individuals CLR** |  |  |  |
| *Time Post Dose + Janssen* | *0.00* | *2* | ***<0.001\*\*\**** |
| Vaccine Type + Time Post Dose | 1.18 | 3 | **0.002\*\*** |
| Vaccine Status + Janssen | 1.52 | 2 | **0.002\*\*** |
| Time Post Dose | 2.19 | 1 | **0.001\*\*** |
| Vaccine Status + Vaccine Type | 2.30 | 3 | **0.003\*\*** |
| Booster Status | 2.68 | 1 | **0.002\*\*** |
| Full Model | 3.06 | 4 | **0.004\*\*** |
| Time Post Dose + Booster Status | 3.51 | 2 | **0.004\*\*** |
| Vaccine Type × Time Post Dose | 3.74 | 5 | **0.005\*\*** |
| Time Post Dose × Booster Status | 5.51 | 3 | **0.012\*** |
| Vaccine Type | 6.72 | 2 | **0.021\*** |
| Janssen | 6.92 | 1 | **0.019\*** |

\* p < 0.05, \*\* p < 0.01, \*\*\* p < 0.001

**Table S2.** AIC results for the two Delta analyses. Minimum AIC score was 75.3, and 82.5 with corresponding log-likelihoods of -36.6 and -34.2 for 55 and 66 pairs, respectively.

| Model | Delta AIC | K | p-value |
| --- | --- | --- | --- |
| **1) Conditional Logistic** |  |  |  |
| **Regression (CLR)** |  |  |  |
| Vaccination Status | 0.00 | 1 | 0.085 |
| Vaccine Type | 1.05 | 3 | 0.116 |
| Time Post Dose | 1.66 | 2 | 0.191 |
| Full Model | 2.50 | 5 | 0.132 |
| Vaccine Type × Time Post Dose | 6.16 | 7 | 0.267 |
| **2) No-age matching CLR** |  |  |  |
| Vaccine Type × Age<sup>1</sup> | 0.00 | 7 | **0.002\*\*** |
| Vaccine Type | 0.02 | 3 | **0.002\*\*** |
| Vaccine Type + Age | 1.99 | 4 | **0.005\*\*** |
| Vaccination Status | 3.91 | 1 | **0.008\*\*** |
| Time Post Dose × Age | 4.68 | 5 | **0.014\*** |
| Full Model<sup>2</sup> | 5.32 | 6 | **0.015\*** |
| Vaccination Status × Age | 5.97 | 3 | **0.029\*** |
| Age | 9.75 | 1 | 0.261 |

<sup>1</sup> Model did not converge
<sup>2</sup> Model converged before all parameters evaluated, some individual parameter estimates should not be used for inference.

**Fig. S1.**
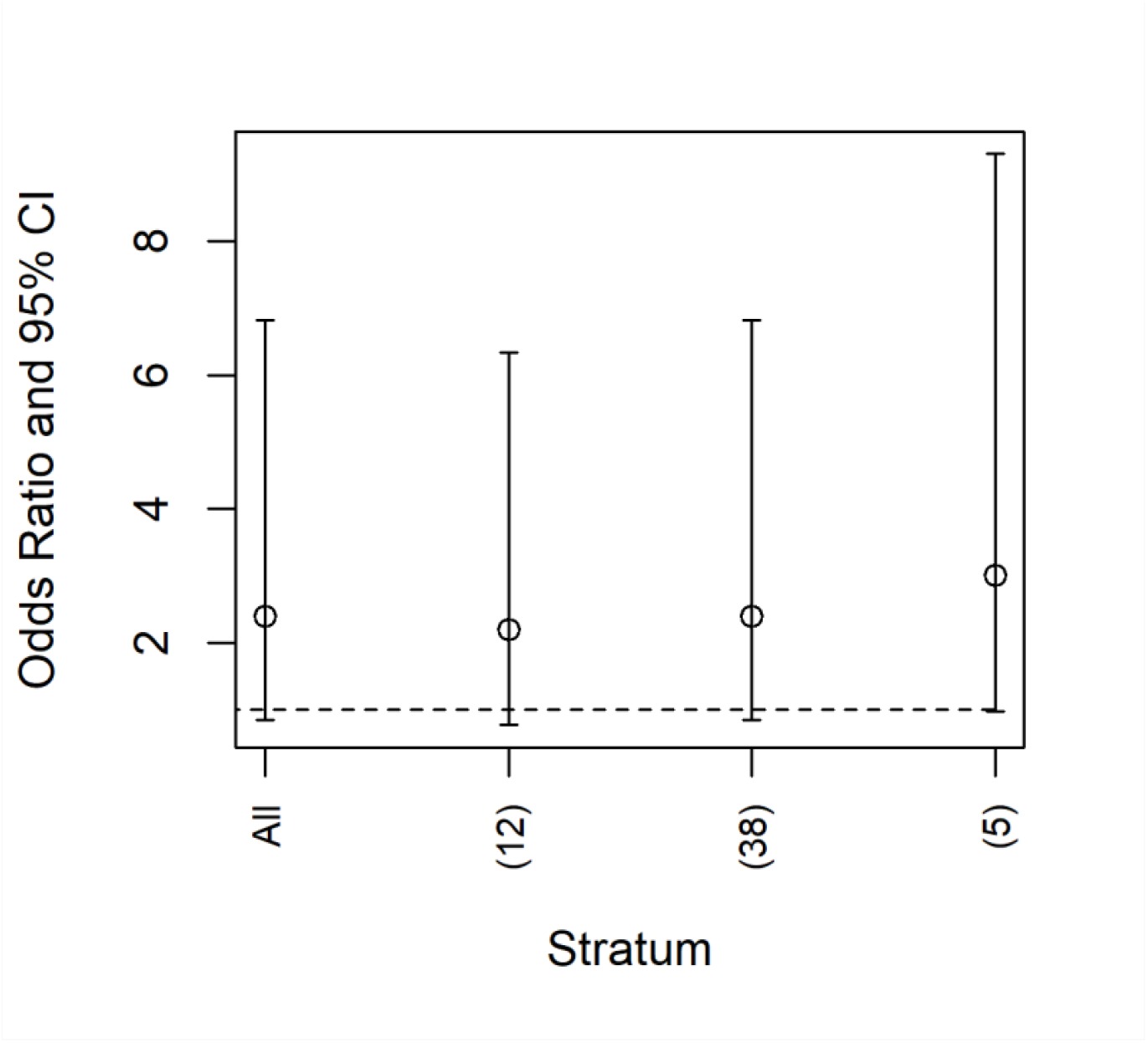
‘All’ denotes the model fit including all pairs. The other entries show the model estimated odds ratio and confidence interval if a single pair was removed. If the removal of two pairs had the same estimated effect size, these data points were grouped together, with the number of pairs with similar effects given on the x-axis. For example, the right-most point shows that there were 5 pairs with similar effect sizes. If any one of those pairs were removed, the confidence interval would be approximately at 1. If two of these pairs were removed, the results would likely be statistically significant. That said, there is no scientific basis for removing these pairs from the analysis. The key point is that the statistical significance, but not the median estimate of the odds ratio, was sensitive to the matching process.

